# Automatic detection of n-degree family members

**DOI:** 10.1101/2025.05.16.25327749

**Authors:** Emil M. Pedersen, Jette Steinbach, Carsten B. Pedersen, Andrew Schork, Morten Krebs, Bjarni J. Vilhjálmsson, Florian Privé

## Abstract

**Summary:** Research in the familial aggregation of diseases and traits utilise information on probands, and their relevant health information enriched with similar information for their family members of interest. The genealogy is typically generated from trio information in registers and biobanks. However it can be tedious and error prone to identify family members other than first-degree relatives. Here, we present a graph-based approach to effectively identify family members of arbitrary degree of relatedness, as well as the means to attach any desired information to each individual for downstream analysis and a function to efficiently calculate a kinship matrix for the identified family members and convert identified family members from a graph back into trio information.

**Availability and Implementation:** The R package where these new functionalities are implemented is available on GitHub (https://github.com/EmilMiP/LTFHPlus) and on CRAN (https://cran.r-project.org/web/packages/LTFHPlus/index.html).

## Introduction

Manually constructing family trees for individuals in large biobanks, population registers, or other study populations is tedious and error-prone. In the field of psychiatry, disorders tend to cluster within families, a finding that generalises to many health outcomes^1,2^. Although genealogy is widely studied, no automated method for inferring family trees up to a given degree of relatedness and linking their information exists. Indeed, only limited resources have recently been dedicated to the analysis of family data and pedigrees in combination with the currently popular genotype data. When looking for methods to construct family trees from trio information (the personal identifier for the target individual, its mother and father), two R packages caught our attention; Kinship2^3^ and FamAgg^4^.

Kinship2 provides methods that can handle family data with a pedigree object. One of the functions included in the package is *makefamid*; a function that can identify families from pedigree information. However, *makefamid* always returns the largest possible family tree and is not intended to find subtrees (see **Figure 1**). For example, Athalandis et al. identified a family of 5,396,661 individuals in the Danish registers using functions similar to *makefamid*^*5*^. Therefore, when considering population based registers such as Danish^6,7^ or Finnish^8^ registers, Kinship2 cannot be used to find family trees of specific degrees of relatedness.

**Figure 1.**
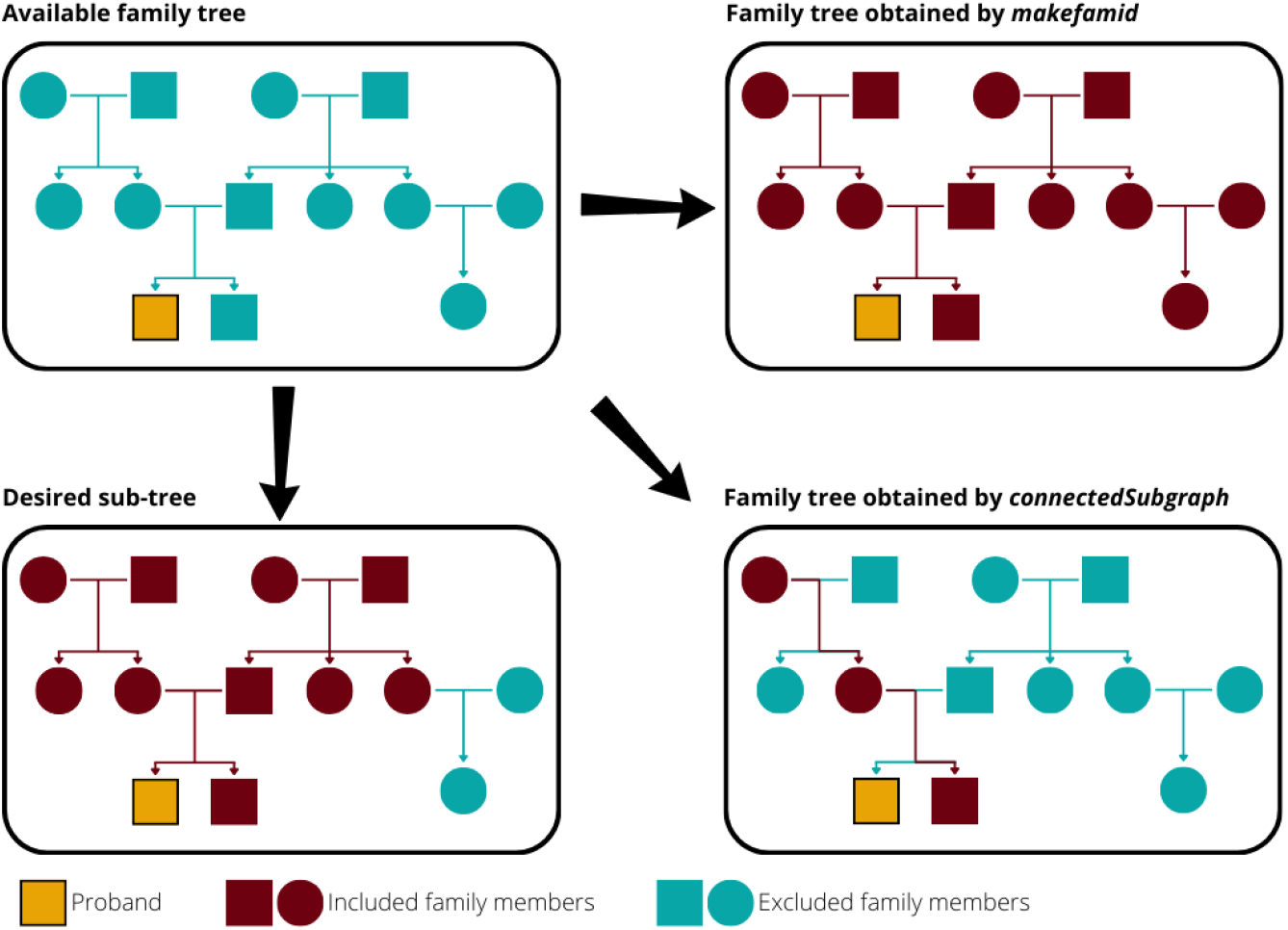
Illustration of the number of family members identified by different R functions.

The function *connectedSubgraph*, included in the package FamAgg^4^, can identify subgraphs in larger family trees represented by a graph. However, *connectedSubgraph* always identifies the smallest possible subgraph that connects a given set of individuals. By doing so, the resulting family tree will ignore any family members that are unnecessary to connect two individuals (see **Figure 1**). Hence, *connectedSubgraph* is too restrictive for our purpose and cannot be used to obtain family trees with all family members of a given degree of relatedness either.

This means that family trees including family members of a pre-specified degree of relatedness have to be constructed manually; a process that is not only time-consuming but also significantly error prone for higher degrees of relatedness. Methods that can generate family trees automatically and efficiently from variables that are already readily available are therefore required.

Here, we designed a new function called *prepare_graph* that is now part of the R package LTFHPlus^9^. *prepare_graph* constructs a (directed) graph from population level input data and attaches additional information to each individual. Using functions from the R package igraph, manipulations can then be performed to this graph to create neighbourhood graphs, i.e. efficiently identify all family members of degree n and closer. A kinship matrix can also be efficiently constructed from a (sub)graph with an additional function *get_kinship* that is also included in LTFHPlus. Graphs can also be converted back into trio information with the function *graph_to_trio*, which makes it possible to utilize properties of existing packages on the identified families, such as plotting with Kinship2.

## Implementation

We implement the function *prepare_graph* that constructs a directed graph from three variables; the personal ID for the target individual along with the personal IDs for the mother and father. This function utilises the R package igraph, which allows for very efficient construction and manipulation of graphs with millions of nodes and edges. In short, the directed graph is obtained through the following steps:

1. Data formatting and quality control, i.e.,
  ∘ Ensure ids are strings
  ∘ Substituting missing values with NAs
  ∘ Conversion to data format used by igraph
2. Identifying parental links
3. Adding edges between full siblings
4. Identifying and adding individuals with no relatives (i.e. isolated nodes)
5. Adding additional individual information (i.e. attaching attributes to nodes)

Once the full population graph is constructed, the function *make_neighbourhood_graph* (alias *make_ego_graph*) from the igraph package can be used to identify all family members of degree n. Given the proband ID (name of a node) or a list of proband IDs, a single n-degree neighbourhood graph is created for each proband. Each neighbourhood graph is centred around the proband, and a proband can also be included as an n-degree relative in the neighbourhood graph for a different proband. Once the desired sub-graph(s) have been obtained, additional manipulations can be performed in order to add, extract, or modify attributes for each identified node in the neighbourhood graph (which corresponds to identifying family members), or to add or remove nodes and edges (which corresponds to adding or removing family members or their relations from the (sub)graph).

In addition, we introduce a second new function in LTFHPlus, *get_kinship*, that calculates the kinship matrix for all individuals present in a (neighbourhood) graph. The kinship matrix is based on the distance between nodes (individuals) in the graph, and it is calculated as 0.5 ^*d*(*i,j*)^ × *C*, where *d*(*i, j*) is the shortest distance between individual *i* and *j* through a most recent common ancestor node, and *C* is a constant. The variable *C* defaults to one, but may represent variables such as the heritability of a phenotype. Zygosity of twins is not considered and, in small families, the shortest path through the most recent common ancestor yields comparable kinship value as inbreeding-aware estimates.

Finally, we also introduce a function called *graph_to_trio*, which takes a graph and reconstructs the trio information used to create it. This step requires sex as an attribute in the graph, however it allows for identification of families of arbitrary degrees and to convert the identified families back into a format that is usable by existing pedigree packages. In the trio format, the identified families can be used in existing pedigree packages, such as the pedigree plotting with Kinship2.

## Usage

The automatic identification of family members presented here has three benefits; remove the potential for errors when identifying family members, the preservation of individual-level family member information, and the construction of kinship matrices. The function *prepare_graph* preserves information for each single family member during the construction of family trees of a suitable degree. Preserving information is helpful when the family trees are used to obtain family-based variables, such as binary family history indicators or family liabilities^10,11^. It also allows for the storage of other parameters on an individual family member level. The second advantage of the approach presented here is the efficient construction of kinship matrices for all identified families. Kinship matrices are applied in many settings; for example, they can be used to account for degrees of relatedness in mixed models^12^, and to construct covariance matrices for liability threshold models conditional on family history^11^. Hence, *get_kinship* can increase efficiency in a broad spectrum of applications.

In particular, we want to highlight a specific application; the estimation of family (genetic) liabilities. The efficient estimation of family (genetic) liabilities is the main purpose of the LTFHPlus package^11,13^. Until now, the identification and construction of family trees and kinship matrices was a major hurdle. But with the automatic identification of family members and the preservation of individual family member information, we have now removed this barrier, minimised the potential for errors, and reduced computation time.

## Conclusion

The new function *prepare_graph* that we added to the R package LTFHPlus efficiently constructs a graph that represents large family trees from trio information. Combined with well established functions such as *make_neighbourhood_graph* from the igraph R package, family trees of arbitrary degrees of relatedness can now be obtained automatically, accurately, and efficiently. Furthermore, we added the function *get_kinship* that can be used to construct kinship matrices for mixed models or the estimation of family (genetic) liabilities and allow for conversion back into trio format for identified families with *graph_to_trio*. We expect these functions to greatly facilitate the continued adoption of family-based analyses.

## Conflicts of interest

BJV is a member of the scientific advisory board for Allelica. The other authors have no conflicts of interest.

## Data Availability

Software is the product of this application note.
The LTFHPlus package can be downloaded from GitHub at github.com/EmilMiP/LTFHPlus or installed directly from CRAN.
Vignettes for usage with example data can be found on the associated pkgdown website (see github).

## Acknowledgement

We thank the Novo Nordisk Foundation for funding for the generation and utilisation of the Danish Multi Generation Register.

## Code availability

The LTFHPlus package can be downloaded from GitHub at github.com/EmilMiP/LTFHPlus or installed directly from CRAN.

## Notes

### Funding Statement

This work was supported by Danish
Data Science Academy, which is funded by the Novo Nordisk Foundation (NNF21SA0069429)

